# Estimated effectiveness of vaccines and extended half-life monoclonal antibodies against respiratory syncytial virus (RSV) hospitalizations in young children

**DOI:** 10.1101/2022.04.19.22272855

**Authors:** Zhe Zheng, Daniel M. Weinberger, Virginia E. Pitzer

**Author notes:** Corresponding Author: Zhe Zheng, Department of Epidemiology of Microbial Diseases, Yale University, 60 College Street, New Haven, Connecticut, USA.

## Abstract

Several vaccines and extended half-life monoclonal antibodies (mAbs) against RSV infection have shown promising progress in clinical trials. Aiming to project the impact of various prevention strategies against RSV hospitalizations in young children, we applied age-structured transmission models to evaluate prevention strategies including maternal immunization, live-attenuated vaccines, and long-lasting mAbs. Our results suggest that maternal immunization and long-lasting mAbs are highly effective in preventing RSV hospitalizations in infants under 6 months of age, averting more than half of RSV hospitalizations in neonates. Live-attenuated vaccines could reduce RSV hospitalizations in vaccinated age groups and are also predicted to have a modest effect in unvaccinated age groups because of disruptions to transmission. A seasonal vaccination program at the country level at most provides a minor advantage regarding efficiency. Our findings highlight the substantial public health impact that upcoming RSV prevention strategies may provide.

## Introduction

Respiratory syncytial virus (RSV) is the leading cause of acute lower respiratory tract infections in infants under 1 year of age globally [1]. Currently, no vaccines or antivirals are available for the prevention and treatment of RSV. The sole pharmaceutical prevention strategy is a monoclonal antibody with a short half-life [2, 3]. However, the high cost and the short duration of this monoclonal antibody limit its use to high-risk infants in high-income countries [4]. Prevention strategies that benefit the general infant population are urgently needed.

Over 40 RSV prophylactic candidates are in pre-clinical or clinical trials [5]. Among them, live-attenuated vaccines, long-lasting monoclonal antibodies (mAbs), and maternal vaccines aim to protect infant populations [5]. Phase 3 clinical trials demonstrate that long-lasting mAbs effectively prevent medically attended RSV-associated lower respiratory tract infections (LRTIs) and hospitalizations throughout the RSV season in healthy preterm, late preterm, and term infants [6-8]. At the same time, phase 2b clinical trials of RSV pre-fusion F protein nanoparticle vaccination in pregnant women suggest that maternal immunization can prevent RSV-associated medically significant LRTIs [9-11]. What remains unclear is the expected effectiveness and vaccine impact of different prevention strategies across age groups, especially with different implementation strategies and levels of coverage.

Several studies have evaluated the potential impact of immunization and prevention strategies for RSV in different settings [12-19]. However, the impact of local variations in RSV epidemic dynamics on the predicted effectiveness of seasonal immunization strategies has not yet been investigated. Moreover, little is currently known about the direct and overall effects of different immunization strategies, which may affect the impact of immunization strategies through herd immunity. Incorporating data from various transmission settings and disentangling direct effects from overall effects, our study set out to estimate the potential impact of three RSV prevention strategies that aim to protect pediatric populations and to identify the key factors that affect vaccine impact.

In this study, we assessed the potential impact of live-attenuated vaccines, long-lasting monoclonal antibodies, and maternal vaccines by modifying a previously published transmission dynamic model of RSV and layering on various prevention strategies [20]. The transmission model was validated against state-specific, age-stratified inpatient data from the United States (US) to account for geographical variations in RSV epidemics. We also conducted a sensitivity analysis to identify key drivers of uncertainty that can be informed by future trials and post-implementation studies.

## Materials and Methods

### Data sources

Individual-level hospital discharge data were obtained from the State Inpatient Databases of the Healthcare Cost and Utilization Project, maintained by the Agency for Healthcare Research and Quality (purchased through the HCUP Central Distributor) [21]. The data covered July 2005 to June 2014 for three states (New York, New Jersey, Washington) and from July 2003 to June 2011 for California. Variables included age at admission (in months for children under 5 years of age and in years otherwise), International Classification of Diseases Ninth Revision (ICD-9) coded diagnoses (multiple fields), and the calendar month and year of hospital admission. Hospitalization was defined as due to RSV if any of the discharge diagnostic codes included 079.6 (RSV), 466.11 (bronchiolitis due to RSV), or 480.1 (pneumonia due to RSV), based on ICD-9. We initialized the transmission dynamic model in 1981 using a total population size equal to the population in the four states in 1980, which was obtained from the 1981 U.S Census Report [22]. We assumed the population age structure of each state remained stable over time and was equal to the age structure for 2010 [23]. We stratified the population into 13 age categories: infants younger than 2 months, 2-3 months, 4-5 months, 6-7 months, 8-9 months, 10-11 months, 1 year, 2-4 years, 5-9 years, 10-19 years, 20-39 years, 40-59 years, and ≥60 years. Birth rate by year and state was obtained from the Centers for Disease Control and Prevention vital statistics [24]. We assumed individuals aged into the next age group exponentially, with the rate equal to the inverse of the length of the age class. We adjusted the net rate of immigration/emigration and death to produce a rate of population growth similar to the observed growth.

### Transmission dynamic models

We extended a previously published age-stratified RSV transmission model [20, 25]. This model assumes newborn infants are protected against RSV infections because they acquire neutralizing antibodies transplacentally from their mothers and/or have few contacts outside the household. With time, transplacentally-acquired antibodies and cocooning effects wane, and infants become susceptible to infection. Following each infection, individuals gain partial immunity that lowers both their susceptibility to subsequent infections and the duration and infectiousness of subsequent infections (see Figure 1). The risk of lower respiratory disease depends on both the number of previous infections and age at infection in the model [20, 25]. We assume frequency-dependent age-specific contact patterns, which were obtained from previous studies that projected contact patterns for the United States [26]. Since RSV epidemics are highly seasonal and vary across states [20], we included state-specific seasonality terms that account for the amplitude and timing of seasonal variation in the transmission rate of RSV.

**Figure 1.**
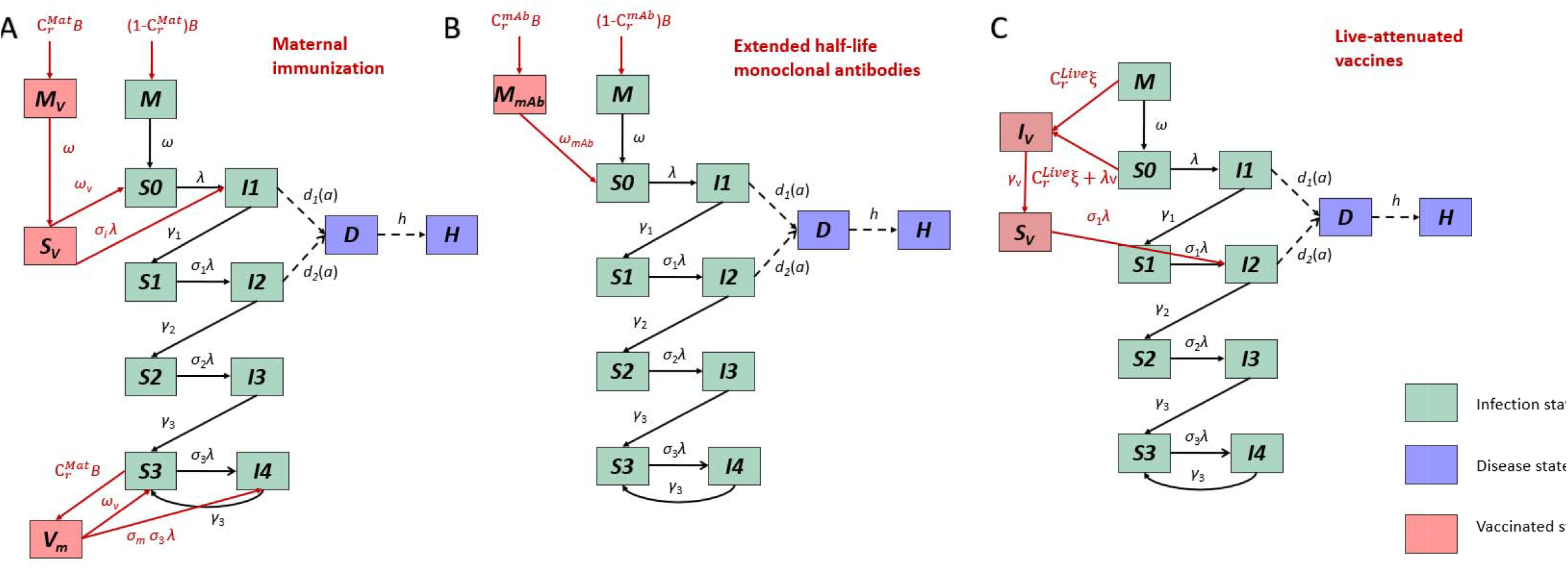
Transmission model structure and immunization strategies. (A) the maternal immunization strategy, (B) extended half-life monoclonal antibodies, and (C) live-attenuated vaccines. The green compartments represent RSV transmission dynamics. Compartments *M* stand for infants who are protected by maternally derived immunity. Compartments *S* stand for susceptible status. The subscripts of *S* compartments represent the number of previous infections. Compartments *I* stand for the infected and infectious status. The subscripts of *I* compartments indicate whether current infection is the first, second, third or subsequent infections. The purple compartments are the observational-level disease states, where *D* stands for lower respiratory tract diseases and *H* stands for being hospitalized. The pink compartments are immunized states. In panel A, represents infants who are born to vaccinated mothers and are fully protected by maternally derived immunity. After the innate maternal immunity wanes, infants who are born to vaccinated mothers become susceptible () to RSV infection with a reduced risk. represents the vaccinated mothers. In panel B, represents infants who receive long-lasting mAbs at birth. In panel C, *I*_*ν*_ represents the newly vaccinated infants who can shed vaccine viruses. After the acute vaccine-shedding period, vaccinated infants gain partial immunity and become less susceptible to RSV infection (*S*_*ν*_).

The majority of the parameters used in the transmission models were fixed based on data from cohort studies conducted in the US and Kenya (Table 1). We estimated a few key parameters that could potentially affect vaccine effectiveness by fitting transmission dynamic models to the hospitalization data from New York, New Jersey, Washington, and California. The estimated parameters included the per capita probability of transmission given contact between an infectious and susceptible individual, the amplitude of seasonality, the seasonal offset, the waning rate of innate maternal immunity, and the reporting fraction (i.e., the probability that a hospitalization caused by RSV is coded as such in the patient record).

**Table 1.** Model parameters.

| Parameter description | Symbol | Parameter value or prior value | Reference | Note |
| --- | --- | --- | --- | --- |
| Transmission dynamic models |  |  |  |  |
| Duration of infectiousness |  |  |  |  |
| First infection | $1/\gamma_1$ | 10 days | | |
| Second infection | $1/\gamma_2$ | 7 days | | |
| Subsequent infection | $1/\gamma_3$ | 5 days | | |
| Relative risk of infection following |  |  |  |  |
| First infection | $\sigma_1$ | 0.76 | [54-57] | |
| Second infection | $\sigma_2$ | 0.6 | | |
| Subsequent infection | $\sigma_3$ | 0.4 | | |
| Relative infectiousness |  |  |  |  |
| Second infections | $\rho_1$ | 0.75 | [54, 55, 58] | |
| Subsequent infections | $\rho_2$ | 0.51 | | |
| Proportion of RSV infections leading to hospitalization |  |  |  |  |
| First infection | | | [58-64] | The probability of hospitalization given infection was estimated as the product of probability of hospitalization given lower respiratory tract infections, probability of lower respiratory tract infections given symptomatic infection ($I_s$), and probability of symptoms given infection: $\Pr(hosp\vert I) = \Pr(hosp\vert LRI) * \Pr(LRI\vert I_s) * \Pr(I_s\vert I)$<br>We estimated the age-specific probability by fitting polynomial regressions to reported aggregate probabilities for 3-month or 6-month age groups |
| 0-2 months old | $h_{p,0-2}$ | 0.082 | | |
| 3-5 months old | $h_{p,3-5}$ | 0.048 | | |
| 6-8 months old | $h_{p,6-8}$ | 0.027 | | |
| 9-11 months old | $h_{p,9-11}$ | | | |
| 1-2 years old | $h_{p,1}$ | | | |
| 2-4 years old | $h_{p,2}$ | | | |
| $\geq 5$ years old | $h_{p,adults}$ | 0.001 | | |
| Second infection | $h_{s,a}$ | $0.4 * h_{p,a}$ | [55] | |
| Third+ infection | $h_{t,a}$ | 0 | [65] | |
| Waning rate of maternal immunity and cocooning effects (1/months) | $\omega$ | $logN(-1, 0.6)$ | [30-33, 66] | |
| Transmission parameter* | $q$ | $N(3, 1)$ | [25] | Truncated at (1, 5) |
| Amplitude of seasonality | $\alpha$ | $N(0.2, 0.05)$ | [25] | |
| Timing of seasonality | $\phi$ | $N(3.4, 1)$ | [25] | Truncated at (0, $2\pi$) |
| Reporting fraction | $\theta$ | $beta(2, 2)$ | [25] | |
| **Vaccine efficacy parameters** |  |  |  |  |
| Relative risk reduction in infants | $\sigma_i$ | 84.7%<br>(50.0%-97.6%) | [9, 28, 29] | Maternal immunization |
| Relative risk reduction in mothers | $\sigma_m$ | 90.0%<br>(80.0%-100.0%) | [9, 45] | Maternal immunization |
| The duration of vaccine-induced protection | $1/\omega_v$ | 5 months | [9, 30-33] | Maternal immunization |
| The duration of protection of extended half-life monoclonal antibodies | $1/\omega_{mAb}$ | 275 days<br>(150 days - 400 days) | [6, 35, 36] | Extended half-life monoclonal antibodies |
| The probability of seroconversion | $\xi$ | 90.0%<br>(80.0%-100.0%) | [38, 39] | Live-attenuated vaccines |
| Duration of shedding of vaccine virus | $1/\gamma_v$ | 5 days | [38, 39] | Live-attenuated vaccines |
| **Coverage parameters** |  |  |  |  |
| Ideal coverage | $C_i$ | 85.0%-95.0% | | |
| **Realistic coverage** |  |  |  |  |
| Maternal immunization | $C_r^{Mat}$ | 53%-70% | [34] | |
| Extended half-life monoclonal antibodies | $C_r^{mAb}$ | 70%-82% | [37] | |
| Live-attenuated vaccines | $C_r^{Live}$ | 70%-86% | [37] | |

\*The basic reproductive number ( $R_0$ ) was estimated from $R_0 = \frac{\det(\beta_{a,k})}{\gamma_1} = \frac{\det(qC_{a,k})}{\gamma_1}$ , using the next-generation matrix method; the transmission parameter $q$ was fitted to the data, $C_{a,k}$ is the contact matrix scaled by the proportion of the population within each age class.

The model fitting process had two steps. We initially seeded the transmission dynamic model with one infectious individual in each age group except for infants under 6 months in July 1981 and used a burn-in period of 24 years in New York, New Jersey, Washington, and 22 years in California to reach quasi-equilibrium. We first used maximum likelihood to fit the model to the observed number of hospitalizations for each state. We then used the maximum likelihood estimates of the number of individuals in each infection state in each age group in July 2005 or July 2003 to initialize the model and refit the transmission dynamic model using Bayesian inference with a gradient-based Markov chain Monte Carlo (MCMC) algorithm. The Bayesian inference allowed us to explore the full parameter space, while the outputs from the maximum likelihood estimation provided a reasonable starting point. The likelihood was calculated by assuming the number of hospitalizations in the entire population in each calendar month was Poisson-distributed with a mean equal to the model-predicted number of hospitalizations, and that the observed age distribution was multinomial-distributed with probabilities equal to the model-predicted distribution of RSV hospitalizations in each age group in children under 5 years in both fitting processes. The prior distribution for each parameter was set as weakly informative (Table 1 and Figure S2). For each state, we sampled 2000 times from 4 chains from the joint posterior distribution of model parameters using the STAN, gradient-based sampling techniques, in R version 4.0.2. We assessed convergence with R-hat, pair plots, and trace plots (Figure S1) [27].

### Modeling three immunization strategies

#### (1) F-protein-based vaccines for pregnant women

We assumed successful maternal vaccination increases the level of transplacentally acquired immunity among infants born to vaccinated mothers and thus reduces the risk of infection in infants after the innate maternal immunity wanes. The lower bound of the relative risk reduction was set at 50% because vaccine candidates are unlikely to be approved if the vaccine efficacy is lower than 50% [28, 29]. The mean and upper bound of relative risk reduction was based on the vaccine efficacy estimates from the exploratory analysis of Phase 2b Pfizer RSVpreF maternal vaccine, which suggested an 84.7% (95% CI 21.6%-97.6%) vaccine efficacy against medically attended RSV lower respiratory tract illness in infants up to 6 months of age. The duration of vaccine-induced protection was assumed to be 5 months because innate maternal immunity was estimated to last for approximately one month [30-33]. We assumed the vaccine may provide additional immunity to pregnant women beyond the natural immunity they already acquired, reducing their risk of getting RSV infection and further protecting their neonates and children through disruptions to transmission. The realistic coverage of maternal immunization was informed by the coverage of maternal influenza vaccination (53%-70%) [34]. To test whether model structure affects effectiveness estimates, we did a sensitivity analysis using a model structure similar to the model structure used for the extended half-life mAbs (see Supplementary documents). We also tested a shorter duration of vaccine-induced protection that lasted for 90 days and a lower risk of infection in vaccinated mothers to see how it affects the estimates of effectiveness (Table S3).

#### (2) Extended half-life monoclonal antibodies for newborns

We assumed that after successful administration of extended half-life monoclonal antibodies at birth, immunized infants will have prolonged immunity against RSV infection. Efficacy is determined by the waning rate of prophylaxis, ω_mab_; we assumed that the average duration of protection (1/ω_mab_) was 275 days (95% CI 150-400 days) [6, 35, 36]. After this prolonged immunity against RSV wanes, immunized infants will become susceptible to RSV infection and will have the same risk of infection as unimmunized infants. The realistic coverage of extended half-life monoclonal antibodies at birth was assumed to be the same as the coverage of hepatitis B vaccine birth dose (70%-82%) [37],

#### (3) Live-attenuated vaccines for seronegative infants

We assumed live-attenuated vaccines will target infants 2-3 months of age with a single dose, as suggested in the overview of pediatric vaccine development plans from World Health Organization workshops [38, 39]. We assumed a successful live-attenuated vaccination (with seroconversion probability ξ set at 0.9 (95% CI 0.8-1)) is comparable to natural infection, since live-attenuated vaccines can induce both humoral and cellular immune responses [38]. After the administration, vaccinated infants can shed vaccine viruses and “infect” susceptible seronegative infants with the vaccine strain, thereby conferring contact immunity [40, 41]. After the acute vaccine-shedding period, vaccinated infants gain partial immunity and become less susceptible to RSV infection. If vaccinated infants are infected with wild-type RSV virus, we assume they have a lower chance of developing severe RSV disease, which was set at 0.76, compared with non-vaccinated infants. The realistic coverage of live-attenuated RSV vaccines was informed by the coverage of rotavirus vaccine (70%-86%) [37].

As there is potential for a booster dose of live-attenuated vaccine in infants, we also tested a delivery strategy that included one booster dose for infants aged 4-5 months as a sensitivity analysis (see Supplementary documents). We assumed a booster dose would also induce both humoral and cellular immune responses comparable to an additional natural infection.

### Effectiveness calculations

We simulated the impact of each intervention strategy for 8 years following implementation. To incorporate uncertainty in the transmission model parameters, we combined the posterior distributions of the model parameters from all four US states. We used Latin hypercube sampling to jointly sample 1000 times from the uncertainty distributions for the transmission model parameters and the efficacy parameters of each prevention strategy. To directly compare the different prevention strategies, we also considered a “high coverage” scenario in which coverage was sampled uniformly from the range 85-95%.

The overall effectiveness of prevention strategies against RSV was measured as the percentage reduction in RSV hospitalizations after implementation compared to the predicted number of RSV hospitalizations with no prevention strategies. We also compared the RSV hospitalization incidence per 100,000 people in each age group after vaccine introduction for 6 years with high coverage to the no prevention scenario. By definition, the direct effect of an intervention is the difference in disease incidence between vaccinated and unvaccinated individuals when all other quantities (in particular, the transmission rate in the population) are comparable [42, 43]. Therefore, to calculate the expected RSV attack rate attributable to the direct effect of vaccination only, we estimated the RSV hospitalization incidence per 100,000 people when holding force of infection (i.e. the per-susceptible transmission rate) at the same level as in an unvaccinated population. The indirect effect can then be estimated as the difference between the overall effect and the direct effect. To measure per-dose efficiency, we divided the overall effectiveness by the total doses given in each prevention strategy.

We compared year-round and seasonal vaccination strategies. The seasonal vaccination strategy was informed by current national recommendations for RSV prophylaxis in the United States, which starts on November 1 and lasts for 5 months for most states. For maternal immunization, we assumed seasonal vaccination will target the infants who are born between November 1 and March 31. For long-lasting monoclonal antibodies, we assumed infants under 6 months who are born outside of the RSV season will be immunized one time on November 1, right before RSV season; infants who are born between November 1 and March 31 will be immunized at birth. For live-attenuated vaccines, we assumed infants aged 2-9 months will be vaccinated on November 1; younger infants will be immunized at 2 months of age between November 1 and March 31. We also evaluated alternative seasonal vaccination strategies that immunize newborns or infants at 2 months of age (1) between November and March or (2) between September and March.

### Variable importance

We used random forest analysis to assess variable importance, since it performs better with non-linear and correlated parameters compared to traditional approaches. Variable importance was measured by the conditional permutation importance method, which measures the prediction error on the out-of-sample portion of the data before and after permuting each predictor variable. This method produces less bias when predictors are correlated compared with the node impurity method or any unconditional permutation importance measure [44]. We explored a wide uncertainty of coverage 40-95% to evaluate the relative importance of coverage in each prevention strategy, along with the uncertainty in the other model parameters. The relative importance was rescaled to be between 0 and 100 to make it comparable across the three prevention strategies.

## Results

### RSV hospitalizations before immunization

The transmission dynamic model reproduces the number of monthly RSV hospitalizations and the detailed age distribution of RSV incidence in each state (Figure S3-6). The state-specific fitted parameters capture the notable variations in the observed timing, seasonal amplitude, and age distribution of RSV epidemics across the four states (Table S1).

### The overall effectiveness of RSV prevention strategies across age groups over time

With high coverage, maternal immunization and long-lasting mAbs offer comparable protection against RSV hospitalizations in the most vulnerable population, those who are under 6 months of age (Table 2). With realistic coverage, monoclonal antibodies may have a larger overall effect than maternal immunization as the expected uptake of maternal immunization is lower (Table 3). With these two prevention strategies, RSV hospitalizations in children over 1 year of age may slightly increase because they delay the age of the first infection (Figure 2A-B and Table 2).

**Table 2.** Percentage of RSV hospitalizations averted across age groups in children under 5 years of age in the United States with coverage ranging from 85% to 95%. Medians and 95% confidence intervals are displayed.

| **Vaccination Strategy** | **Maternal immunization (%)** | **Monoclonal antibodies (%)** | **Live-attenuated vaccines (%)** |
| --- | --- | --- | --- |
| **0-1 month** | 53 (34, 63) | 58 (31, 76) | 31 (11, 45) |
| **2-3 months** | 40 (25, 49) | 50 (25, 64) | 64 (44, 79) |
| **4-5 months** | 28 (17, 36) | 41 (19, 54) | 65 (50, 77) |
| **6-7 months** | 19 (11, 25) | 32 (14, 45) | 66 (53, 77) |
| **8-9 months** | 11 (6, 16) | 24 (9, 37) | 66 (54, 76) |
| **10-11 months** | 4 (0, 8) | 16 (3, 28) | 65 (54, 76) |
| **1 Yr** | -12 (-25, -4) | -11 (-22, -2) | 61 (51, 72) |
| **2-4 Yrs** | -23 (-56, -7) | -42 (-109, -20) | 50 (39, 63) |
| **<5 Yrs** | 24 (15, 30) | 31 (15, 40) | 53 (39, 64) |

**Table 3.** Total RSV hospitalizations averted per 100,000 people in children under 2 years of age. Medians and 95% confidence intervals are displayed.

| **Vaccination Strategy** | **High year-round coverage** | **High seasonal coverage** | **Realistic year-round coverage** |
| --- | --- | --- | --- |
| **Maternal immunization** | 250 (160, 380) | 120 (80, 150) <sup>+</sup> | 170 (110, 270) |
| **Monoclonal antibodies** | 340 (190, 470) | 290 (1900, 360) <sup>*</sup> | 290 (160, 420) |
| **Live-attenuated vaccines** | 470 (370, 640) | 390 (290, 470) <sup>*</sup> | 410 (310, 550) |

<sup>+</sup> Covered infants who are born between November and March
<sup>\*</sup> Cover infants at risk between November and March with a one-time catch up before
RSV season

**Figure 2.**
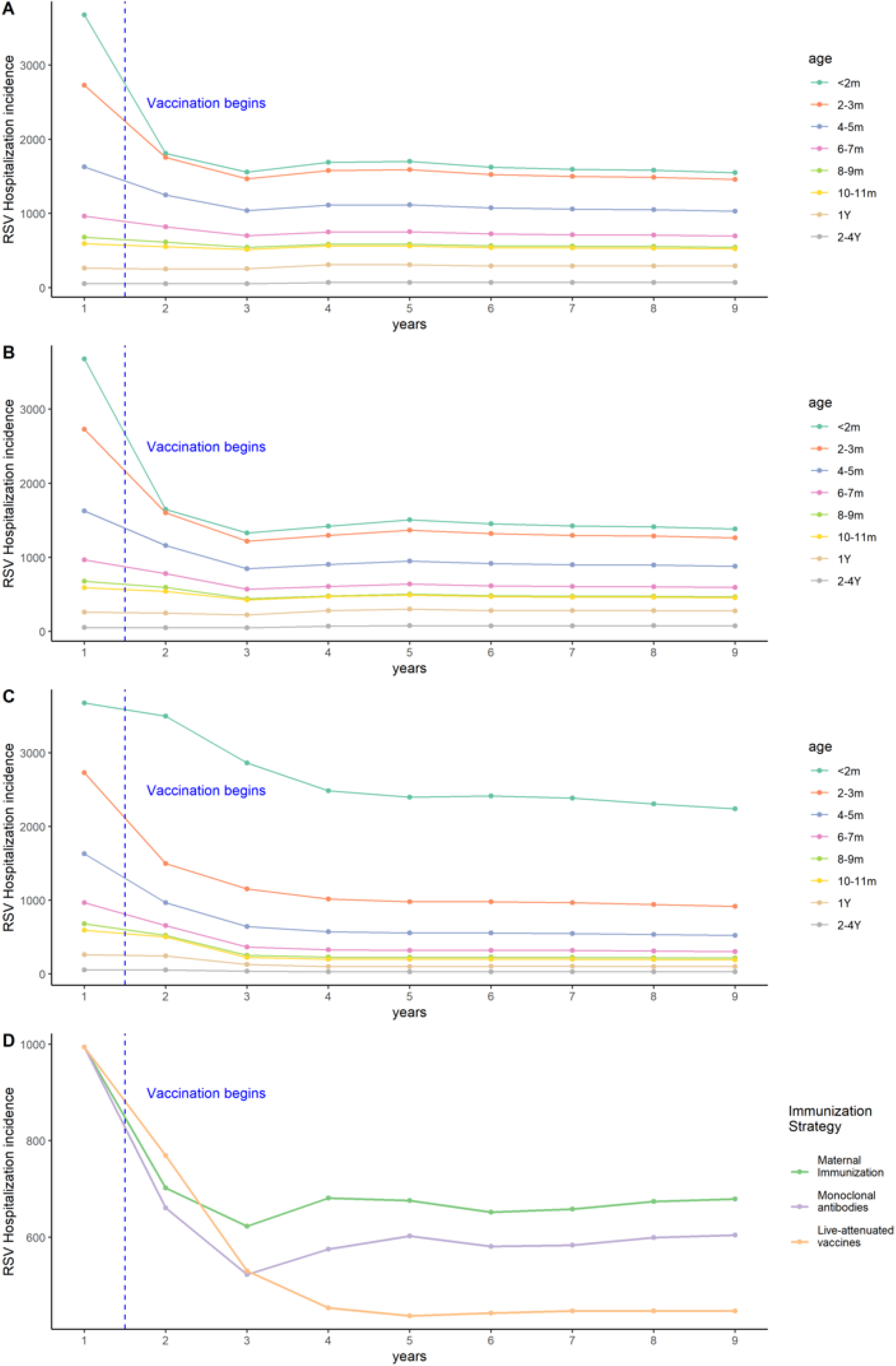
RSV hospitalization incidence per 100,000 people across age groups over time in three RSV prevention strategies. Annual RSV hospitalization incidence per 100,000 people across age groups over time for (A) the maternal immunization strategy, (B) extended half-life monoclonal antibodies, and (C) live-attenuated vaccines. (D) Monthly RSV hospitalization incidence per 100,000 people for all infants <2 years of age for each RSV prevention strategy. We assumed 85%-95% coverage. The color lines show the mean RSV hospitalization incidence before and after implementation of RSV prevention strategies in each age group over time.

Live-attenuated vaccines have a strong effect in the target age groups receiving the vaccines, averting more than two-third of RSV-associated hospitalizations (Table 2). There is little effect in younger age groups that are not eligible for the vaccine in the first year that vaccination program begins (Figure 2C). However, a modest effect in the unvaccinated age groups increases over time due to reduced transmission in the population (Figure 2C). Moreover, compared with the other two strategies, live-attenuated vaccines lead to continued decline in RSV hospitalizations in children over 1 year of age (Table 2 and Figure 2C).

### Estimated overall and direct effects

Maternal immunization and long-lasting mAbs provide direct protection to newborn infants immediately after birth (Figure 3A and Figure 3B). The overall effects of maternal immunization and long-lasting mAbs are similar to their direct effects because these strategies are not expected to influence transmission (Figure 3A). The overall effects of live-attenuated vaccines are greater than the direct effects across all age groups. This difference is especially obvious in infants under 2 months of age who are not eligible for receiving vaccines and are not protected by direct effects. (Figure 3C).

**Figure 3.**
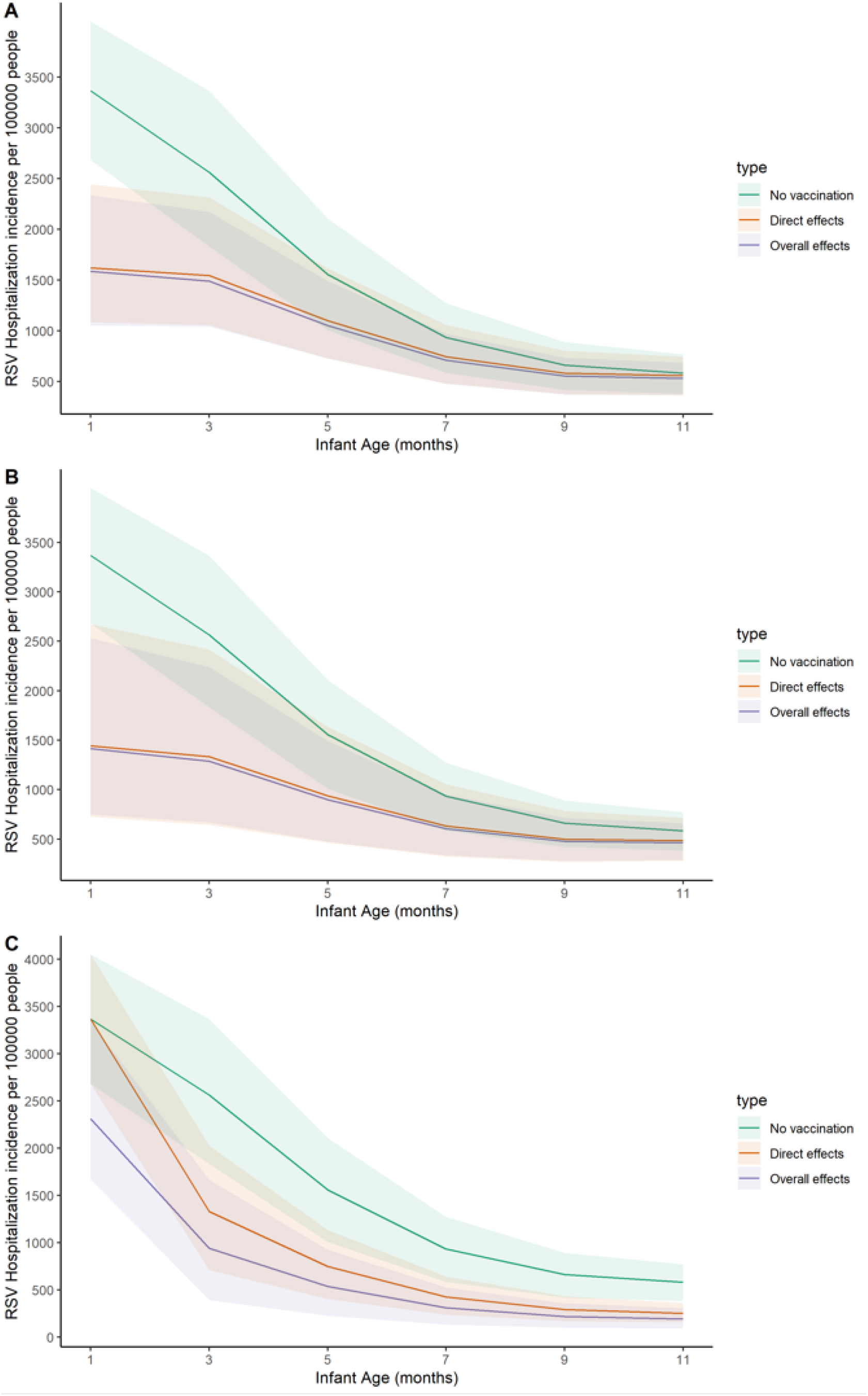
Overall effects and direct effects of three RSV prevention strategies. The model-predicted RSV hospitalization rate per 100,000 people by age is plotted for: (A) maternal immunization, (B) extended half-life monoclonal antibodies, and (C) live-attenuated vaccines. The green lines show the mean RSV hospitalization incidence assuming no vaccination. The orange lines show the model-predicted direct effects of RSV prevention strategies (i.e. assuming no reduction in RSV transmission). The purple lines show the overall effects of RSV prevention strategies, which account for both the direct and indirect effects among the vaccinated age groups and the indirect effects among the unvaccinated age groups. The color shadows show the 95% confidence interval in the same group.

### Estimated effectiveness under different implementation scenarios

While maternal immunization and long-lasting mAbs provide the greatest protection for neonates, live-attenuated vaccines may provide the largest overall effect for populations under 2 years of age (Table 2, Table 3, and Table 4). In the scenario with high year-round coverage, live-attenuated vaccines could avert 470 (370, 640) RSV-associated hospitalizations per 100,000 people compared with 250 (160, 380) RSV-associated hospitalizations averted by maternal immunization and 340 (190, 470) by long-lasting mAbs.

**Table 4.** RSV hospitalizations averted per 1000 doses. Medians and 95% confidence intervals are displayed.

| **Vaccination Strategy** | **Year-round coverage** | **Nov-Mar seasonal coverage** | **Sep-Mar seasonal coverage** | **Routine + catch-up** |
| --- | --- | --- | --- | --- |
| **Maternal immunization** | 5.1 (3.2, 7.6) | 6.0 (3.8, 7.5) | 6.6 (4.2, 8.3) | NA |
| **Monoclonal antibodies** | 6.7 (3.9, 9.2) | 7.2 (3.3, 10.1) | 8.1 (4.2, 10.2) | 7.3 (4.9, 9.1) |
| **Live-attenuated vaccines** | 11.9 (8.8, 15.5) | 12.1 (8.6, 15.1) | 12.6 (9.3, 15.9) | 9.0 (6.6, 10.7) |

Seasonal immunization strategies have lower overall effectiveness compared with year-round immunization strategies, which is most evident for the maternal immunization strategy (Table 3). Seasonal maternal immunization averts 120 (80, 150) RSV-associated hospitalizations per 100,000 people, which is half of the hospitalizations averted under the year-round coverage strategy. Seasonal immunization with a one-time catch-up before the RSV season yields a slightly lower overall effectiveness compared with year-round immunization strategies.

Despite the lower impact on rates of hospitalization, seasonal immunization strategies may provide slightly higher per-dose effectiveness (Table 4). For example, under year-round vaccination, maternal immunization was predicted to avert 5.1 (3.2, 7.6) RSV hospitalizations per-1000 doses. Under seasonal maternal vaccination from September to March, the number of RSV hospitalizations averted per 1000 doses was estimated to be 6.6 (4.2, 8.3). The coverage scenario did not affect per-dose effectiveness. While the relative efficiency of seasonal immunization is high for infants under 6 months of age, the relative efficiency of seasonal immunization is below 1 for infants aged 6-11 months (Table S2).

### Sensitivity analysis

The model parameters that determine the vaccine efficacy are most important in estimating the per-dose effectiveness of each immunization strategy (Figure 4). These include the relative risk reduction of RSV infection in infants born to vaccinated mothers, the duration of protection by monoclonal antibodies, and the immunogenicity of live-attenuated RSV vaccines. The reduced risk of infection in mothers has little impact on the per-dose effectiveness of maternal immunization, even after considering a wider possible range (Figure 4A). For long-lasting mAbs, the duration of innate maternal immunity may also be associated with per-dose effectiveness (Figure 4B). For live-attenuated vaccines, the reporting fraction and transmission parameter are also associated with estimates of per-dose effectiveness (Figure 4C).

**Figure 4.**
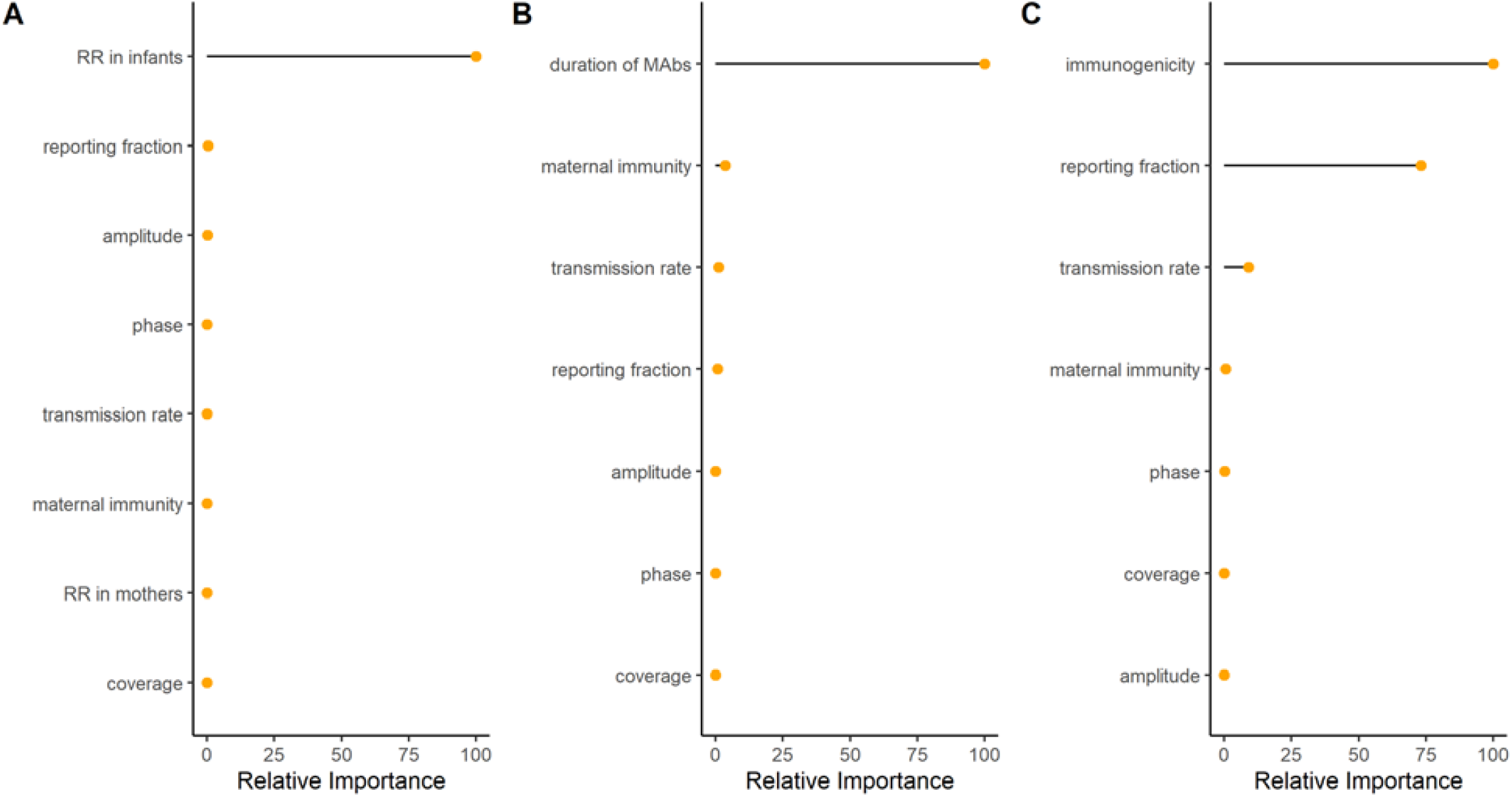
Relative importance of variables for the predicted per-dose effectiveness of the three RSV prevention strategies. The relative importance of variables for model predictions of the per-dose effectiveness of (A) maternal immunization, (B) extended half-life monoclonal antibodies, and (C) live-attenuated vaccine are plotted. From top to bottom shows the most important factor in determining vaccine effectiveness to the least important factor in each prevention strategy.

## Discussion

As multiple RSV prevention strategies targeting pediatric populations are in the final stages of clinical development and testing, it is important to understand the potential population impacts of these prevention strategies. Applying compartmental models, we set out to estimate and compare the overall effectiveness of three different prevention strategies across age groups over time in the United States. Our results suggest that maternal immunization and long-lasting mAbs protect the most vulnerable, those who are under 6 months of age, but they will not provide substantial additional indirect effects for the pediatric population. Live-attenuated vaccines have a lower predicted impact initially, particularly for infants less than 2 months of age, but offer additional benefits by interrupting RSV transmission. In addition, live-attenuated vaccines reduce RSV hospitalizations in children over 1 year of age whereas the other two strategies lead to a slight increase in hospitalization incidence in older children as they delay the first time of infection.

Our study estimated higher overall effectiveness of maternal immunization compared with previous studies [13-16, 18]. The main difference comes from the assumptions regarding vaccine efficacy (see Supplements). Most of the previous studies used efficacy estimates from the phase 3 clinical trial of the Novavax maternal RSV vaccine candidate or assumed a protective duration of 90 days [45]. However, as the clinical trial failed to meet the prespecified success criterion, it is unlikely that a government agency like U.S. Food and Drug Administration will approve this product. Furthermore, a 90-day average protective duration generates a vaccine efficacy estimate below 50% compared to the estimated current level of maternally-derived protection. Our study updated the efficacy estimates based on the latest progression in clinical trials. Our results also suggested that the reduced risk of infection in mothers has little effect in determining the per-dose effectiveness of maternal immunization. The effect of maternal immunization in reducing transmission is predicted to be modest. These results are in line with previous studies [14, 46]. Since our model did not consider household structure, the effects of maternal immunization in reducing transmission could be underestimated. However, clinical trials suggested that the risk of RSV disease in vaccinated pregnant women was not different from those who were unvaccinated.

Our results suggested that long-lasting mAbs have high effectiveness against RSV hospitalizations in infants under 6 months of age. As our models are based on inpatient data of general populations, the high effectiveness indicates the potential of long-lasting mAbs to be administered as universal prophylaxis for every infant, especially if the price is comparable to a vaccine [35]. Our results indicated that the effectiveness of long-lasting mAbs is similar to that of maternal immunization across all age groups. Thus, these two prevention programs are likely to be interchangeable. However, long-lasting mAbs have unique advantages in preventing RSV-associated hospitalizations in preterm infants, as our sensitivity analysis suggested that the duration of innate maternal immunity is associated with the overall effectiveness estimates of long-lasting mAbs. Long-lasting mAbs will be a preferable prevention strategy for preterm infants, since preterm labor curtails the total amount of transplacentally acquired antibodies in infants and thus leads to a decreased efficacy of maternal immunization [47].

Live-attenuated vaccines were estimated to be the most effective immunization strategy in children under 2 years of age. Although live-attenuated vaccines do not directly protect the youngest infants, as their immature immune and respiratory system makes them ineligible for receiving live-attenuated RSV vaccines, these vaccines were predicted to provide indirect protection to newborns by decreasing RSV transmission in the entire population. While live-attenuated vaccines were the best strategy for reducing hospitalization incidence overall among children under 2 years of age, this strategy alone predicted the lowest reduction in RSV hospitalizations among neonates, who are at highest risk of severe outcomes. Combining live-attenuated vaccines targeting infants older than 6 months with either maternal immunization or long-lasting mAbs at birth could potentially provide good protection across the pediatric age spectrum.

One counterintuitive finding of our model predictions for live-attenuated vaccines was that a strategy with one-time catch-up seasonal vaccination had the lowest per-dose effectiveness compared with other year-round or seasonal vaccination strategies. This is because with a campaign targeting infants aged 2-9 months, some vaccines will be given to RSV seropositive infants, who do not respond to the live-attenuated vaccines [48-51]. Thus, identifying a level of coverage that induces herd immunity while minimizing the wasted doses in seropositive infants and young children could be an interesting topic for future research. If further development of these vaccines leads to increased immunogenicity in seropositive infants and young children, both the overall effectiveness and per-dose effectiveness of live-attenuated vaccines will increase.

In contrast to an earlier finding that suggested a seasonal RSV vaccination strategy was much more efficient than a year-round vaccination strategy [17], our estimates of per-dose effectiveness of the seasonal vaccination strategy were only slightly higher than the year-round vaccination strategy. One major difference between our study and the previous study is that we estimated the per-dose effectiveness in children under 2 years of age, while they looked at infants under 6 months old. We found that although seasonal vaccination was more efficient in infants under 6 months of old, infants aged 6-11 months were not well protected by a seasonal vaccination strategy (Table S2). This is because infants born in late spring and summer, who will not be protected by a seasonal immunization strategy, will encounter their first RSV season when they are over 6 months of age. Furthermore, our study considered state-specific RSV seasonality within the US, while the previous study was done at the national level across low- and middle-income countries. With high spatial variations in RSV timing in the US [20, 52], a country-level seasonal vaccination strategy is unlikely to be efficient [53].

Our study is subject to several limitations. First, the overall effectiveness estimates of our study highly relied on the input efficacy. Although we used the latest efficacy estimates from clinical trials and applied a reasonable lower bound as input parameters, efficacy estimates may change as clinical trials progress. Therefore, our overall effectiveness estimates need to be interpreted with caution. These analyses were not intended to provide a precise point estimate for each strategy. Instead, the focus is on comparison between the three prevention strategies, particularly with respect to the overall effectiveness in different age groups. Second, the numbers for RSV-associated hospitalizations only reflect a proportion of the “true” disease burden. This limitation will not affect the overall effectiveness, since it is measured as a percentage decrease. However, underreported RSV-associated hospitalizations will lead to underestimates of per-dose effectiveness. This may explain why RSV-associated hospitalizations averted per 1000 infants immunized in clinical trials with active surveillance is twice what we estimated in our study [8]. Third, we did not consider household structure in our analysis, which may lead to underestimated effects of maternal immunization in reducing transmission. Nonetheless, a previous study that considered household structure also suggested that maternal immunization would induce little herd immunity [14]. Fourth, live-attenuated vaccines may have protective effects in seropositive children that we did not account for in our analysis. We assumed that live-attenuated vaccines have no effect on seropositive children because phase I clinical trial data suggested a comparable antibody level in seropositive children before and after vaccination. However, antibody level may not directly translate into vaccine protection. If live-attenuated vaccines confer additional protection among seropositive children, we anticipate a higher overall effectiveness and per-dose effectiveness.

## Conclusions

In this study, we used compartmental models within a Bayesian framework to estimate state-specific transmission parameters and incorporated the uncertainties in transmission settings to estimate the overall effectiveness of three RSV prevention strategies in detailed age groups over time. We predicted that maternal immunization and long-lasting mAbs would be highly effective in infants under 6 months of age, while a live-attenuated vaccine would be the most effective immunization strategy for children under 2 years of age as it was predicted to provide both direct and substantial indirect protection. A seasonal vaccination program at the country level would provide only a slight efficiency advantage over a year-round vaccination program. Our findings join previous research highlighting the substantial public health impact that upcoming RSV prevention strategies may provide.

## Data Availability

The data and code that support the findings of this study are available via GitHub (https://github.com/weinbergerlab/rsv-transmission-model). The hospitalization data are not available publicly but can be obtained upon signing a data use agreement with the Agency for Healthcare Research and Quality.

## Acknowledgements

We thank Dr. Louis Bont for helpful discussions on these analyses.

## Author contributions

ZZ conceived and implemented the study, analyzed the data, and drafted the article. DMW and VEP conceptualized the study and reviewed and revised the manuscript. All authors have seen and approved the final draft of the manuscript.

## Competing interests

VEP has received reimbursement from Merck and Pfizer for travel expenses to Scientific Input Engagements on respiratory syncytial virus. DMW has received consulting fees from Pfizer, Merck, GSK, Affinivax, and Matrivax for work unrelated to this manuscript and is the principal investigator on research grants from Pfizer and Merck on work unrelated to this manuscript. All other authors report no relevant conflicts.

## Funding

Research reported in manuscript was fully supported by the National Institute of Allergy and Infectious Diseases (MIDAS Program) of the National Institutes of Health under award number R01AI137093. The content is solely the responsibility of the authors and does not necessarily represent the official views of the National Institutes of Health.

## Notes

### Author Declarations

Ethics committee/IRB of Yale University waived ethical approval for this work

