## Supplementary document for "Estimated effectiveness of vaccines and extended half-life monoclonal antibodies against respiratory syncytial virus (RSV) hospitalizations in young children"

**Supplementary documents**

**Supplementary methods**

*Transmission dynamic models*

We defined a seasonal age-specific force of infection that varies with time. The force of infection $\lambda_{a}\left( t \right)$ for age group $a$ and time *t* is defined as:

$$\lambda_{a}\left( t \right)=\left( 1+b_{1}cos\left( \frac{2\pi t-\phi}{12} \right) \right)\sum_{k} \beta_{a,k}(I_{1,k}\left( t \right)+\rho_{1}I_{2,k}\left( t \right)+\rho_{2}I_{3,k}\left( t \right){+\rho_{2}I_{4,k}\left( t \right))}/{N_{k}}\left( t \right)$$

It contains three major components: the seasonal transmissibility of RSV, the age-specific transmission parameter, and the transmissibility related to the number of infections. The seasonal dynamic of RSV is represented by $(1+b_{1}cos(\frac{2\pi t-\phi}{12}))$, where $b_{1}$is the amplitude of seasonality in transmission and *ϕ* is the timing of peak transmissibility. The transmission parameter $\beta_{a,k}$ is the product of the per capita probability of transmission given contact between an infectious and a susceptible individual (*q*) and contact rate between age group *k* and age group $a$ ($C_{a,k}$). The age-specific contact patterns were obtained from previous studies that projected the contact patterns to the United States [2, 3]. These two components are multiplied by the number of infectious individuals of age *k* who have been infected one, two, three and four or more times at time $t$: ${(I}_{1,k}\left( t \right)+\rho_{1}I_{2,k}\left( t \right)+\rho_{2}I_{3,k}\left( t \right){+\rho_{2}I_{4,k}\left( t \right))}/{N_{k}}\left( t \right)$, where the relative infectiousness of second and subsequent infections are denoted as $\rho_{1}$ and $\rho_{2}$; the total population of age *k* at time $t$ is denoted as $N_{k}\left( t \right)$. We stratified the population into 13 age groups considering their risk of developing severe RSV diseases and contact patterns: infants younger than 2 months, 2-3 months, 4-5 months, 6-7 months, 8-9 months, 10-11 months, 1 year, 2-4 years, 5-9 years, 10-19 years, 20-39 years, 40-59 years, and ≥60 years.

The transmission dynamic process is link to observational inpatient data. We assume that every infected individual has a probability $h_{i,a}$ of developing severe RSV disease that requires hospitalization, which depends on infection order *i* and age *a*, and a fraction $\theta$ of RSV hospitalizations will be recorded in the inpatient datasets:

$$H_{a}\left( t \right)=\theta*(\lambda_{a}\left( t \right){(S}_{0,a}\left( t \right)h_{p,a}+{\sigma_{1}S}_{1,a}\left( t \right)h_{s,a}+{\sigma_{2}S}_{2,a}\left( t \right)h_{t,a}+{\sigma_{3}S}_{3,a}\left( t \right)h_{t,a}))$$

where $H_{a}\left( t \right)$ is the number of RSV hospitalizations in age group *a* at time *t* and $\lambda_{a}\left( t \right)$ is the force of infection that age group $a$ experiences at time $t$. The fully susceptible individuals of age *a* who have never been infected before are denoted as $S_{0,a}\left( t \right)$. The number of susceptible individuals of age *a* who have been infected one, two, and more times at time $t$ are denoted as $S_{1,a}\left( t \right), S_{2,a}\left( t \right),$ and $S_{3,a}\left( t \right)$, respectively; $\sigma_{1}, \sigma_{2},$ and $\sigma_{3}$ represent the reduced susceptibility to RSV infection following the first, second, and more infections due to the partial immunity gained after each infection. $h_{p,a}$, $h_{s,a}$ and $h_{t,a}$ are the proportion of the first, second, and subsequent infections in age group $a$ that require hospitalizations, respectively.

*Relative efficiency calculation*

To determine the efficiency of seasonal prevention strategies compared to year-round prevention, we calculated the ratio of per-dose effectiveness between a seasonal program and a year-round program in each age group. That is,

$$\frac{\frac{\text{RSV hospitalizations averted in age group} k \text{in a seasonal program}}{\text{number of vaccine doses given in a seasonal program}}}{\frac{\text{RSV hospitalizations averted in age group} k \text{in a year-round program}}{\text{number of vaccine doses given in a year-round program}}}.$$

**Supplementary results**

**Table S1. State-specific estimated (median and 95% credible interval) transmission dynamic model parameters.**

|  | New Jersey | New York | Washington | California |
| --- | --- | --- | --- | --- |
| Basic reproductive number* (*R*_0_) | 10.37 (10.24, 10.52) | 10.16 (10.12, 10.26) | 10.11 (9.96, 10.26) | 9.76 (9.72, 10.04) |
| Timing of seasonality d) | 1.35 (1.34, 1.37) | 1.36 (1.35, 1.37) | 1.44 (1.43, 1.45) | 1. 40 (1.40, 1.41) |
| Amplitude of seasonality (*b*_1_) | 0.20 (0.19, 0.21) | 0.16 (0.16, 0.17) | 0.20 (0.18, 0.21) | 0.25 (0.23, 0.26) |
| Duration of maternal immunity and cocooning effects (1/*ω*30.44,* in days) | 90.84 (78.34, 104.43) | 74.44 (69.02, 80.71) | 41.07 (33.02, 50.70) | 4.62 (3.41, 17.31) |
| Reporting fraction | 0.76 (0.68, 0.87) | 0.95 (0.84, 0.99) | 0.63 (0.53, 0.81) | 0.89 (0.62, 0.97) |

*The basic reproductive number (*R*_0_) was estimated from $R_{0}=\frac{det(\beta_{a,k})}{\gamma_{1}}=\frac{det({qC}_{a,k})}{\gamma_{1}},$ using the next-generation matrix method; the parameter *q* was fitted to the data, $C_{a,k}$ is the contact matrix scaled by the proportion of the population within each age class.

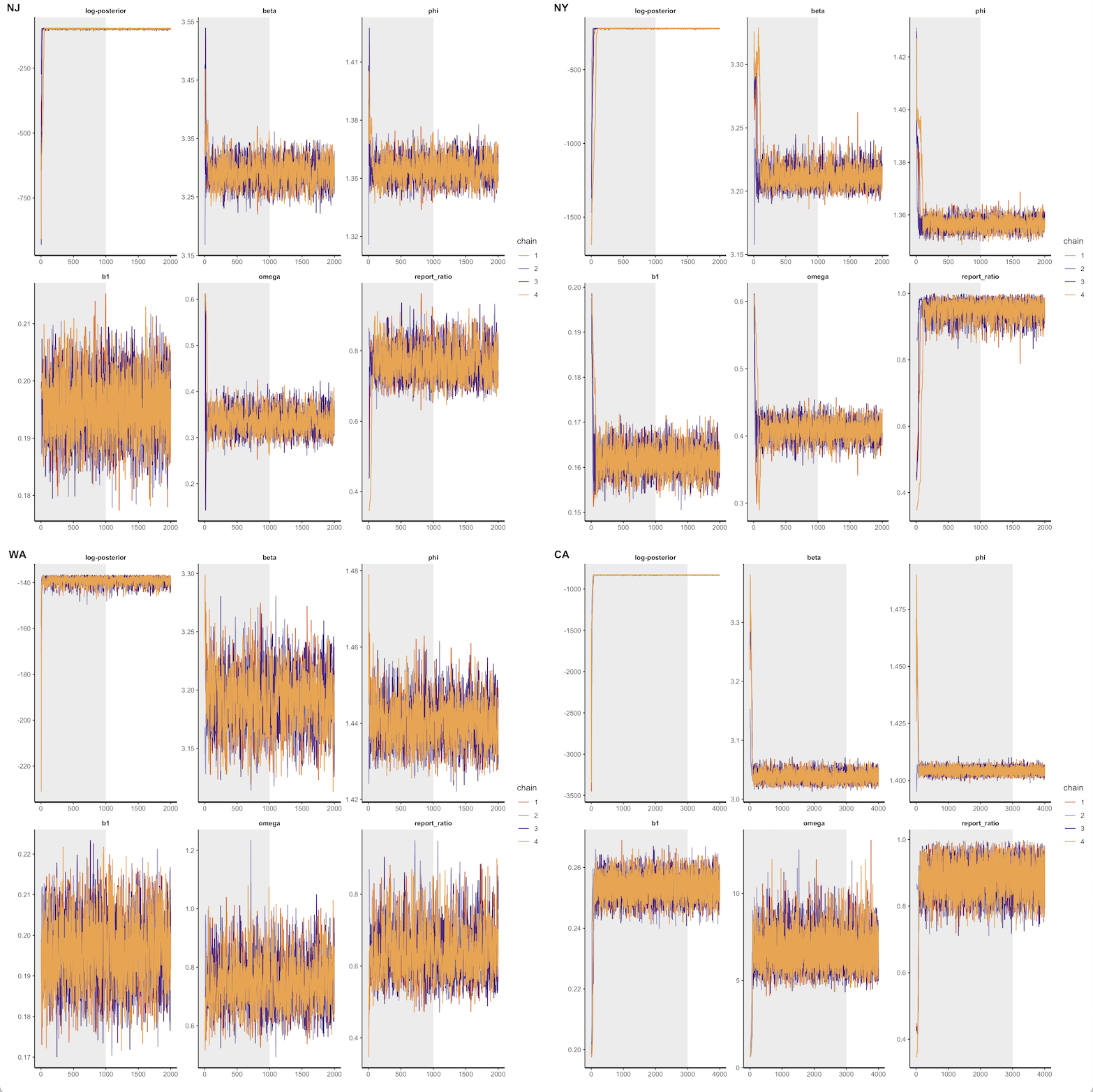

**Figure S1. Parameter trace plots for four states.** Posterior draws of the log posterior model probability and five parameters from New Jersey (NJ), New York (NY), Washington (WA), and California (CA). The grey shaded areas indicate the burn-in period. The color lines indicate four chains of posterior draws. All parameters converged well in all states.

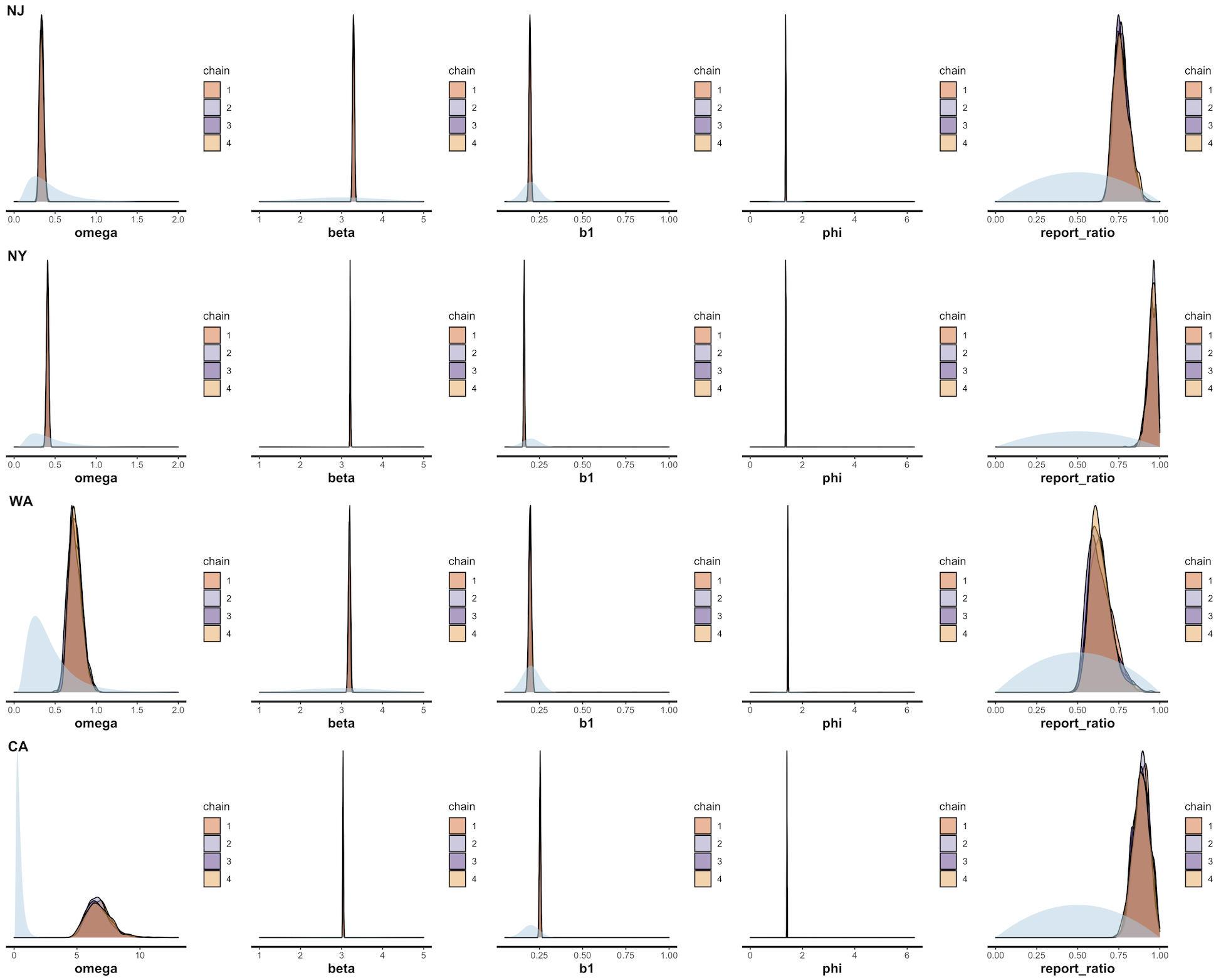
**Figure S2. Prior and posterior distributions of state-specific estimated transmission dynamic model parameters.** The light blue areas represent the weakly-informative prior distribution for each parameter. The color shaded areas represent the posterior distribution of state-specific estimated transmission dynamic model parameters in each chain.

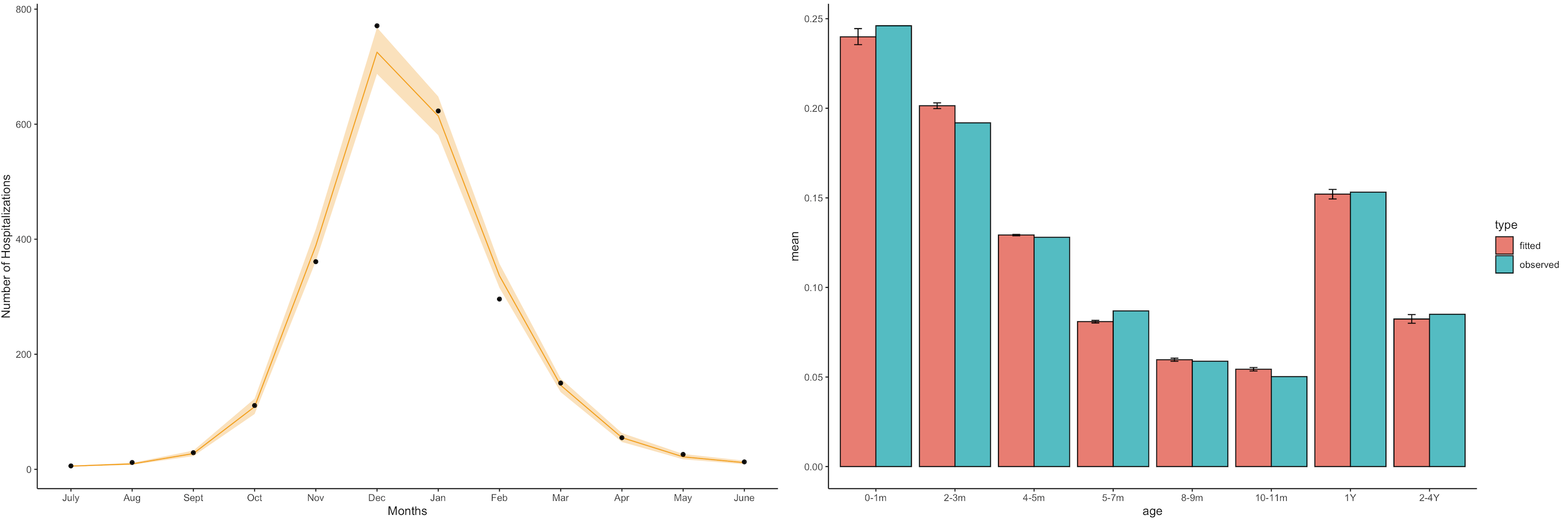

**Figure S3. Model fit to mean number of RSV hospitalization per month and age distribution in New Jersey.** On the left panel, the ICD9-CM coded hospitalization data is represented by the dots. The median of the fitted model is shown in solid yellow line while the shaded area around the line shows the 95% credible interval. On the right panel, proportion of hospitalizations in each age group for the ICD9-CM coded hospitalization data is shown in blue, and the corresponding proportions for the fitted models are shown in red.

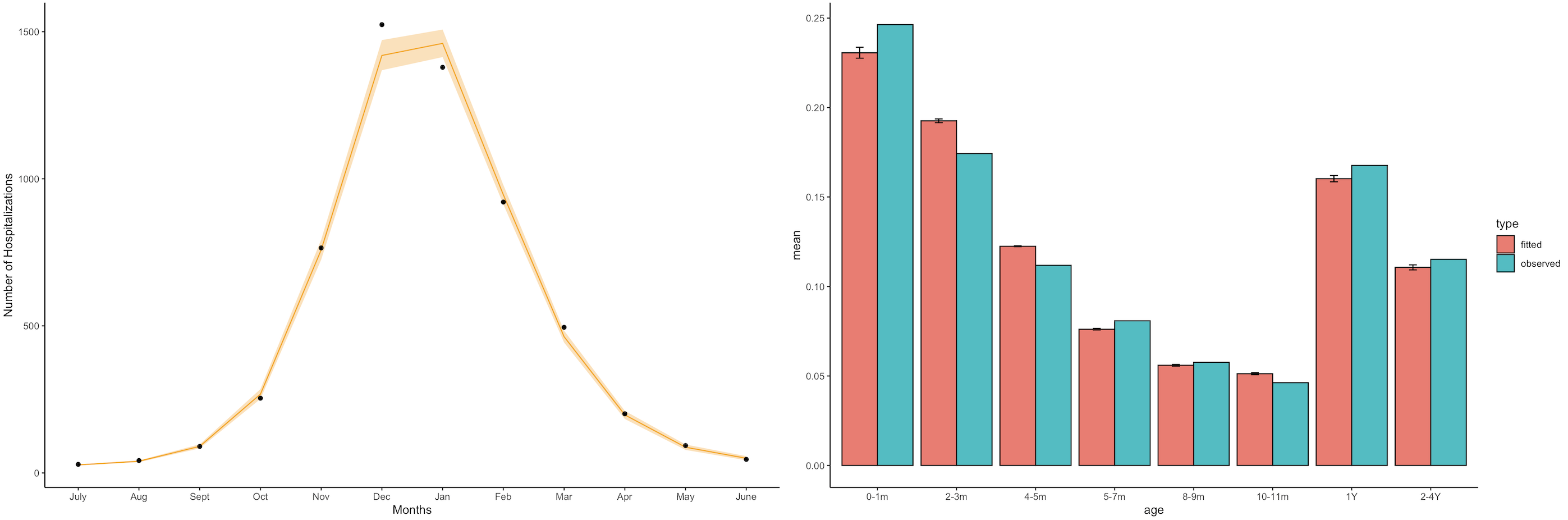

**Figure S4. Model fit to mean number of RSV hospitalization per month and age distribution in New York.** On the left panel, the ICD9-CM coded hospitalization data is represented by the dots. The median of the fitted model is shown in solid yellow line while the shaded area around the line shows the 95% credible interval. On the right panel, proportion of hospitalizations in each age group for the ICD9-CM coded hospitalization data is shown in blue, and the corresponding proportions for the fitted models are shown in red.

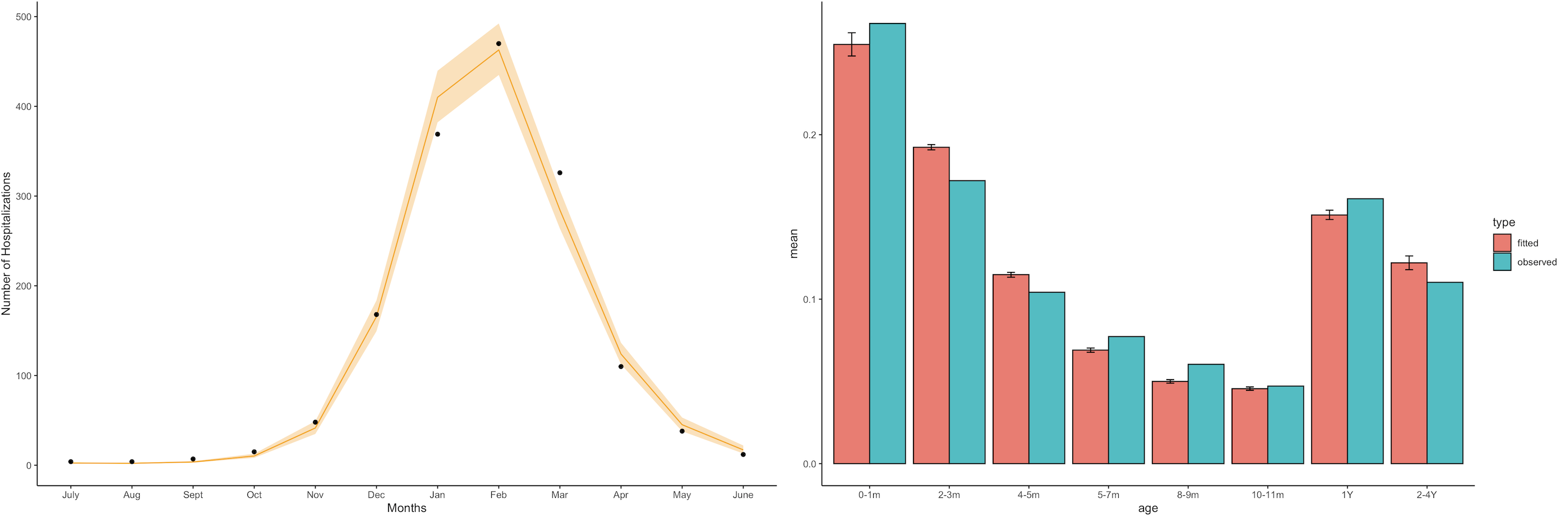

**Figure S5. Model fit to mean number of RSV hospitalization per month and age distribution in Washington.** On the left panel, the ICD9-CM coded hospitalization data is represented by the dots. The median of the fitted model is shown in solid yellow line while the shaded area around the line shows the 95% credible interval. On the right panel, proportion of hospitalizations in each age group for the ICD9-CM coded hospitalization data is shown in blue, and the corresponding proportions for the fitted models are shown in red.

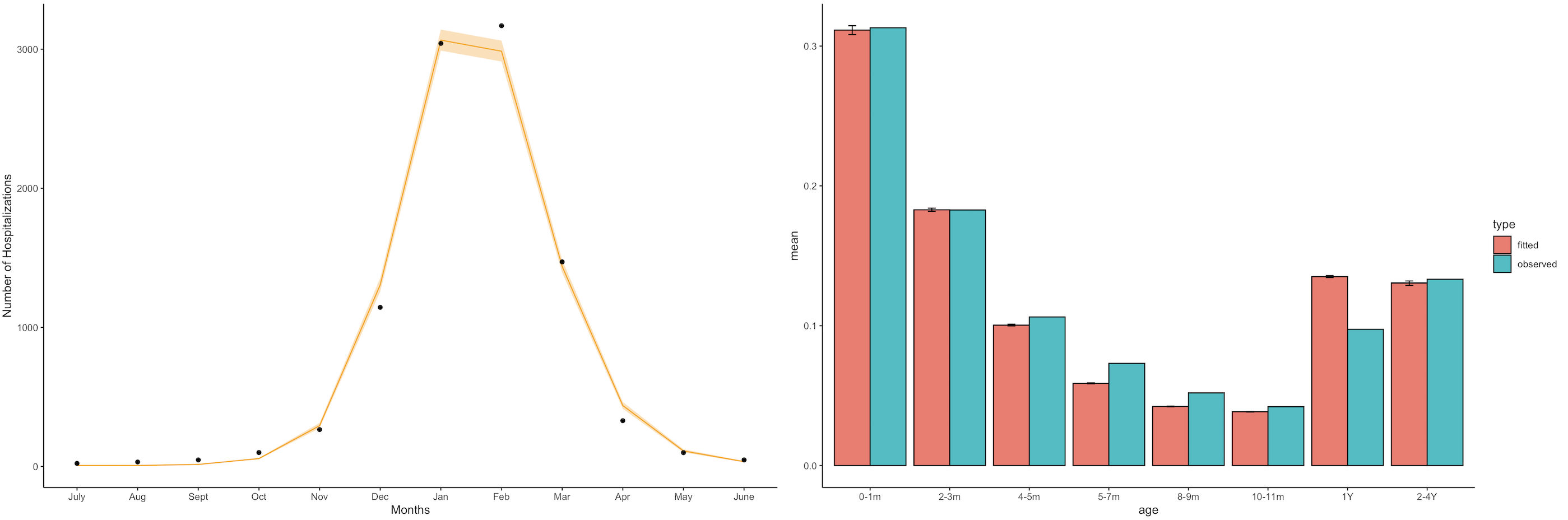

**Figure S6. Model fit to mean number of RSV hospitalization per month and age distribution in California.** On the left panel, the ICD9-CM coded hospitalization data is represented by the dots. The median of the fitted model is shown in solid yellow line while the shaded area around the line shows the 95% credible interval. On the right panel, proportion of hospitalizations in each age group for the ICD9-CM coded hospitalization data is shown in blue, and the corresponding proportions for the fitted models are shown in red.

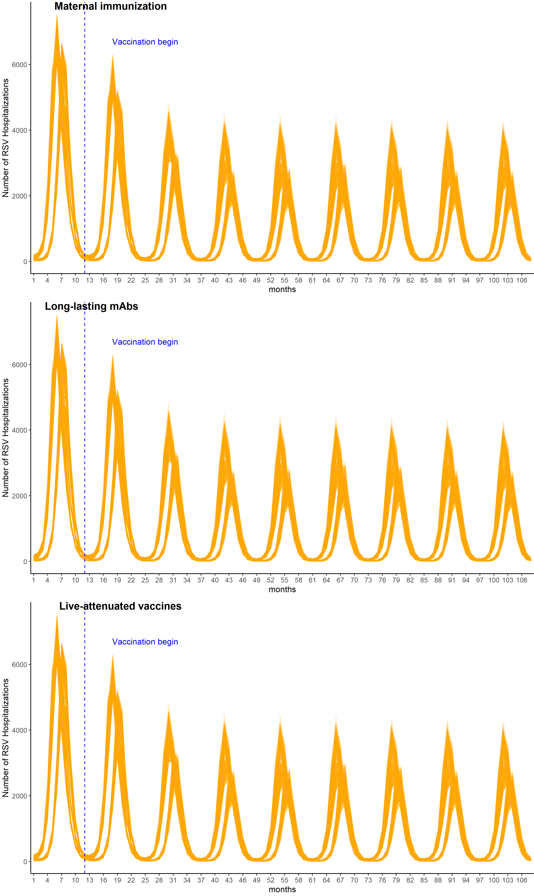

**Figure S7. RSV-associated hospitalizations from each run.** Total number of RSV hospitalizations over time for (A) the maternal immunization strategy, (B) extended half-life monoclonal antibodies, and (C) live-attenuated vaccines. Each line represents the result of one set of parameter combinations.

**Table S2. Relative efficiency between year-round vaccination strategy and Sep-Mar seasonal vaccination strategy.** The number of hospitalizations averted per 1000 doses is shown for each strategy and age group.

| **Vaccination Strategy** | **Maternal immunization** | **Monoclonal antibodies** | **Live-attenuated vaccines** |
| --- | --- | --- | --- |
| ***Age group***  **0-1 month** | 1.45 | 1.37 | 0.92 |
| **2-3 months** | 1.19 | 1.10 | 1.29 |
| **4-5 months** | 0.91 | 0.86 | 1.08 |
| **6-7 months** | 0.67 | 0.70 | 0.94 |
| **8-9 months** | 0.36 | 0.64 | 0.90 |
| **10-11 months** | -2.43 | 0.55 | 0.94 |
| **Overall** | 1.28 | 1.18 | 1.05 |

*Sensitivity analysis*

*Maternal immunization*

To test how model structural assumptions may affect the overall effectiveness estimates of maternal immunization programs, we modified the model structure by assuming maternal immunization provides prolonged passive immunity to newborn infants while also lowering the risk of infection in mothers. We assumed the passive immunity from maternal immunization has an average duration of 1/ $\omega_{\mathrm{mat}}$ = 175 days (95% CI 150-200 days) in newborn infants.

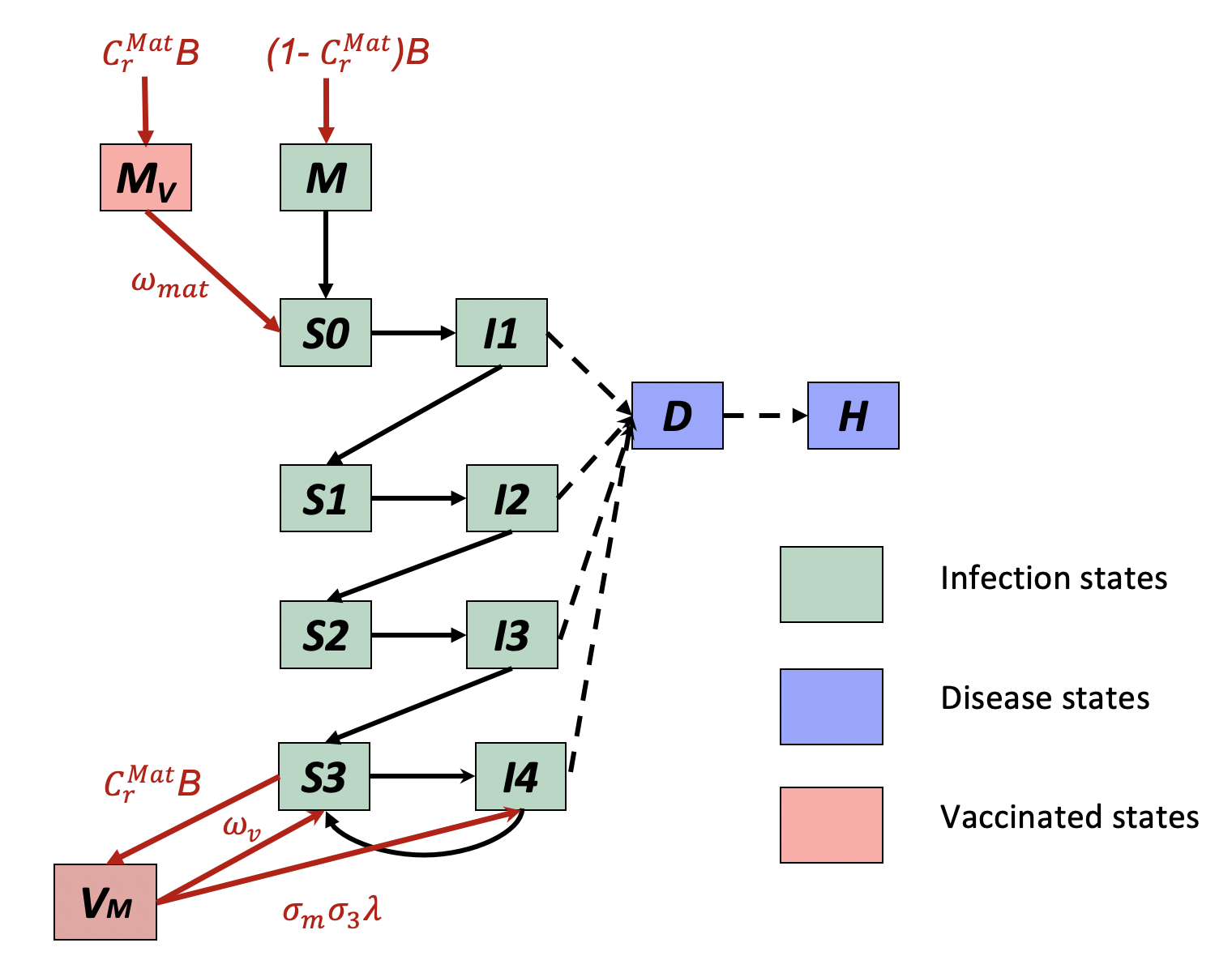

**Figure S8. Alternative model structure of maternal immunization programs.** The green compartments represent RSV transmission dynamics. The purple compartments are the observational level diseases states. The pink compartments are immunized states.

We also tested two other assumptions to see how they affect the estimates of effectiveness, including (1) a shorter duration of vaccine-induced protection that lasts for 90 days and (2) a lower risk of infection in vaccinated mothers compared with unvaccinated mothers that ranges from 0.4 to 0.6.

**Table S3. Comparison of maternal immunization effectiveness under different assumptions.**

| **Vaccination Strategy** | **Original maternal immunization (%)** | **Alternative model structure (%)** | **Shorter duration of vaccine-induced immunity (%)** | **Lower risk of infection in vaccinated mothers (%)** |
| --- | --- | --- | --- | --- |
| **0-1 month** | 53 (34, 63) | 46 (27, 69) | 45 (29, 53) | 54 (36, 64) |
| **2-3 months** | 40 (25, 49) | 37 (21, 53) | 29 (19, 37) | 41 (27, 50) |
| **4-5 months** | 28 (17, 36) | 29 (16, 39) | 18 (10, 25) | 29 (19, 39) |
| **6-7 months** | 19 (11, 25) | 21 (11, 28) | 10 (4, 15) | 20 (12, 28) |
| **8-9 months** | 11 (6, 16) | 13 (7, 19) | 3 (0, 6) | 12 (6, 18) |
| **10-11 months** | 4 (0, 8) | 7 (2, 12) | -3 (-5, 0) | 4 (1, 9) |
| **1 Yr** | -12 (-25, -4) | -8 (-15, -4) | -12 (-24, -7) | -14 (-31, -8) |
| **2-4 Yrs** | -23 (-56, -7) | -23 (-42, -11) | -18 (-41, -8) | -32 (-73, -14) |
| **<5 Yrs** | 24 (15, 30) | 22 (13, 30) | 18 (12, 23) | 25 (16, 31) |

An alternative model structure of maternal immunization yielded similar effectiveness estimates. Shorter duration of maternal immunity leads to a lower vaccine effectiveness, which is an 18% (12%, 23%) reduction in RSV-associated hospitalizations in children under 5 years old (compared with a 24% reduction if the duration is 5 months). Lower risk of RSV infection in vaccinated mothers yielded a marginal increase in effectiveness estimates in children under 5 years of age.

*Live-attenuated vaccines*

We tested the assumption of one booster dose of live-attenuated vaccines for infants aged 4-5 months. We assumed a booster dose would also induce both humoral and cellular immune responses comparable to an additional natural infection.

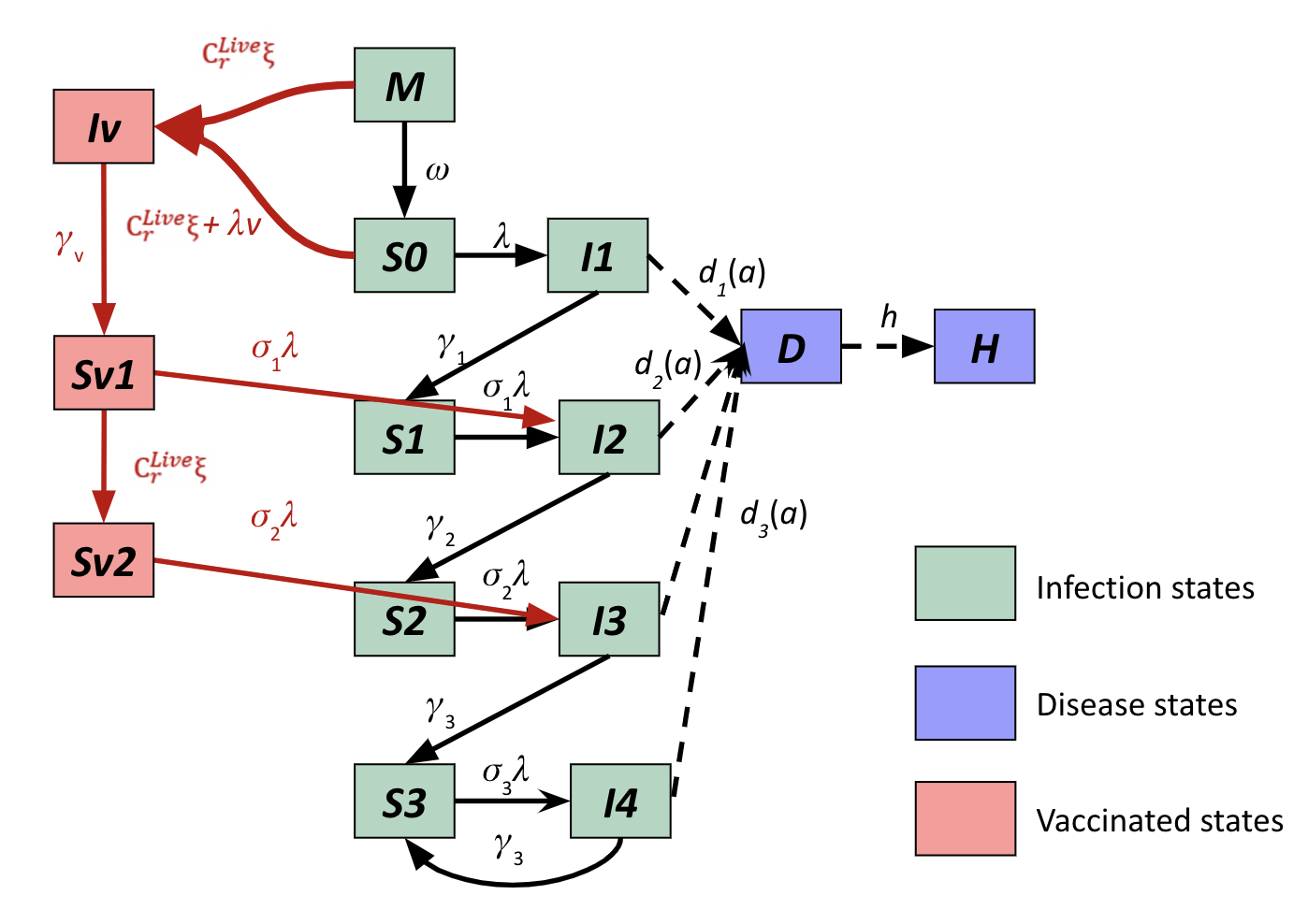

**Figure S9. Alternative model structure of a booster dose of live-attenuated vaccines for infants.** The green compartments represent RSV transmission dynamics. The purple compartments are the observational level diseases states. The pink compartments are immunized states.

The booster dose of the live-attenuated RSV vaccine increased the overall vaccine effectiveness in every age group in children under 5 years of age. Infants aged 4-5 months benefited the most from the live-attenuated booster dose, resulting in an 83% (68%, 94%) reduction in RSV-associated hospitalizations (Table S4). Overall, live-attenuated vaccines with a booster dose will lead to a 67% (49%, 80%) reduction in RSV-associated hospitalizations in children under 5 years old (compared with a 53% reduction without a booster dose).

**Table S4. Comparison of effectiveness of live-attenuated vaccine with and without booster dose.**

| **Vaccination Strategy** | **Live-attenuated vaccines without booster dose (%)** | **Live-attenuated vaccines with booster dose (%)** |
| --- | --- | --- |
| **0-1 month** | 31 (11, 45) | 44 (14, 61) |
| **2-3 months** | 64 (44, 79) | 71 (46, 85) |
| **4-5 months** | 65 (50, 77) | 83 (68, 94) |
| **6-7 months** | 66 (53, 77) | 83 (68, 94) |
| **8-9 months** | 66 (54, 76) | 83 (68, 93) |
| **10-11 months** | 65 (54, 76) | 82 (68, 93) |
| **1 Yr** | 61 (51, 72) | 78 (54, 90) |
| **2-4 Yrs** | 50 (39, 63) | 66 (50, 82) |
| **<5 Yrs** | 53 (39, 64) | 67 (49, 80) |
